# Dynamics of corticocortical brain functional connectivity relevant to therapeutic response to biologics in inflammatory arthritis

**DOI:** 10.1101/2022.05.15.22275083

**Authors:** Nobuya Abe, Kodai Sakiyama, Yuichiro Fujieda, Khin K. Tha, Hisashi Narita, Kohei Karino, Masatoshi Kanda, Michihito Kono, Masaru Kato, Tatsuya Atsumi

**Affiliations:** Department of Rheumatology, Endocrinology and Nephrology, Faculty of Medicine and Graduate School of Medicine, Hokkaido University, Sapporo, Japan; Department of Diagnostic and Interventional Radiology, Hokkaido University Hospital, Sapporo, Japan; Global Centre for Biomedical Science and Engineering, Faculty of Medicine, Hokkaido University, Sapporo, Japan; Department of Psychiatry, Faculty of Medicine, Hokkaido University, Sapporo, Japan; Department of Rheumatology and Clinical Immunology, School of Medicine, Sapporo Medical University, Sapporo, Japan

**Keywords:** resting-state functional magnetic resonance imaging, dynamic functional connectivity, neuroimaging, rheumatoid arthritis, spondyloarthritis, biologics, inflammatory arthritis, disease activity, brain function

## Abstract

Aberrant resting-state static functional connectivity of the brain regions, which could be evaluated by functional magnetic resonance imaging (fMRI), affects clinical courses in inflammatory arthritis (IA) including rheumatoid arthritis and spondyloarthritis. This static methods for assessing brain functional connections would be too simple to estimate the whole picture of resting-state brain function because it fluctuates over time. The effects of resting-state brain connectivity dynamics for clinical course are unknown in patients with IA. Therefore, we aimed to evaluate dynamic functional connectivity for clinical courses of IA in the context of therapeutic responsiveness to biologics using resting-state fMRI data of 64 patients with IA consisting of two cohorts. We determined representative whole-brain dynamic functional connectivity patterns by *k*-means++ cluster analysis, and evaluated the association of their occurrence probability and therapeutic outcomes with biologics. We determined four distinct clusters of dynamic functional connectivity in IA patients. In the first cohort, occurrence probability of the distinct cluster was associated with favorable therapeutic response in disease activity and patients’ global assessment. This finding was validated by the second cohort. The whole-brain functional coordination of the cluster indicated significantly increased corticocortical connectivity, and probabilistically decreased after therapy in treatment-effective patients compared to -ineffective patients. In conclusion, dynamic functional connectivity, in particular, frequent emergence of corticocortical connections was associated with clinical outcomes in patients with IA. The coherence of corticocortical interactions might affect modulation of pain, which would be relevant to therapeutic satisfaction.

**SUMMARY:**

- Effects of resting-state dynamic connectivity on clinical course of inflammatory arthritis regarding therapeutic responsiveness to biologics were assessed by functional magnetic resonance.
- Occurrence probability of corticocortical functional connectivity pattern was associated with favorable therapeutic response in disease activity and patients’ global assessment in inflammatory arthritis.

## INTRODUCTION

The interaction between central nervous system and peripheral system including pain sensation and immunity is of current interest and has been investigated [13]. In autoimmune inflammatory arthritis (IA) such as rheumatoid arthritis (RA) and spondyloarthritis (SpA), disease activity status including patients’ global assessment and systemic inflammation levels are affected by brain function, which could regulate perception and peripheral immunity. Vice versa, brain function is affected by chronic pain and systemic inflammation. Grading the disease activity and deciding the management of IA is sometimes difficult because they may be subjected to patients’ self-assessment of pain, of global health or of disease activity [9]. These subjective parameters would be affected by brain function. Therefore, evaluating individual brain function is possibly useful for assessing the disease activity and determining the therapeutic strategy in patients with IA.

Functional magnetic resonance imaging (fMRI) is one of the neuroimaging methods, which can detect blood oxygen level-dependent (BOLD) signals reflecting functional activity in each brain region, and functional connectivity between brain regions can be calculated using Pearson’s correlation [20]. In IA, previous fMRI studies demonstrated the clinical utility of assessing functional connectivity between brain regions. For instance, higher levels of inflammation are associated with more positive connectivity among the inferior parietal lobule, medial prefrontal cortex, and multiple brain networks, and these patterns of connections predicted the presence of increased fatigue and pain in RA patients [6; 29]. A study reported a significant positive correlation between default-mode network connectivity to the left insula and a continuous measure of the degree of fibromyalgia in RA [5]. Interestingly, RA patients with fibromyalgia showed peripheral inflammation through pronociceptive patterns of brain connectivity between insula and anterior cingulate cortex (ACC) [17]. In patients with chronic pain, previous studies demonstrated that brain functional connectivity could predict clinical course of the disease in terms of pain chronification and pharmacologic response [4; 16]. We have previously demonstrated that functional connectivity between left insular cortex and ACC was associated with therapeutic responsiveness to biologics in patients with IA [1].

The general method for calculating functional connectivity uses the entire BOLD time series derived from a prolonged period of fMRI acquisition to generate a summary correlation, which is termed static functional connectivity. However, brain functional connectivity actually fluctuates over time, indicating that the static methods for functional connections would be too simple to estimate the whole picture of resting-state brain functions in a given individual [24]. To overcome this problem, dynamic functional connectivity matrices were extracted from the acquired fMRI data by the sliding-window approach segmenting the BOLD time series and the calculation of correlations between brain regions [18].

Although the association of resting-state dynamic functional connectivity with clinical outcomes has been evaluated in the realm of chronic pain like fibromyalgia or osteoarthritis [7; 8], it is unknown whether the dynamic functional connectivity is also associated with clinical courses in patients with RA and SpA. Thus, in the current study, we explanatory investigated the association of resting-state dynamic functional connectivity with longitudinal clinical parameters and outcomes in IA patients.

## PATIENTS and METHODS

### Patients

Patients who met 2010 American College of Rheumatology (ACR) RA classification criteria [2], or Assessment of SpondyloArthritis international Society (ASAS) classification criteria for axial and peripheral SpA [26; 27] were selected. Data of patients with IA including RA and SpA from two cohorts, which include 33 patients and 31 patients, respectively, were collected at Hokkaido University Hospital from 2018 to 2019 and 2019 to 2020, respectively (Supplementary Table 1, 2) [1]. These patients needed therapeutic intensification by biological disease modifying anti-rheumatic drugs and were clinically evaluated before and three months after initiation of therapy (Supplementary Table 3, 4). Clinical assessments for RA included a simplified disease activity index (SDAI) [30]. Twenty-eight joints for tender joint count (TJC) and swollen joint count (SJC) which include shoulders, elbows, knees, wrists, and each finger and thumb (metacarpophalangeal or proximal interphalangeal joints). Patient’s global assessment (PGA) and evaluator’s global assessment (EGA) are assessed by a visual analogue scale scored from 0 to 10. Clinical assessments for SpA included Ankylosing Spondylitis Disease Activity Score (ASDAS)-C-reactive protein (CRP), which is calculated by the degrees of back pain and peripheral pain/swelling, morning stiffness duration, PGA and serum level of CRP [21]. The patients were functionally evaluated by the modified Health Assessment Questionnaire (mHAQ) for patients’ functional status in daily living activities (range: 0-3.0) and Fibromyalgia Impact Questionnaire (FIQR) for evaluating the total spectrum of problems related to fibromyalgia. FIQR contains 3 domains regarding (i) function for daily living, (ii) overall impact for accomplishing goals and overwhelmed state, and (iii) symptoms such as pain, fatigue and mental difficulty, graded on a 0–10 numeric scale in each item. Effective response was defined as at least 20% improvement in ACR core set of disease activity measures in RA [12], and ASAS 20 improvement [3]. We obtained written-informed consent for the study and publication from all participants. The study protocol was approved by the Institutional Review Board of Hokkaido University Hospital (reference number: 010-0031, 018-0128 and 018-0222). The present study complied with the Declaration of Helsinki.

### Image acquisition

All brain imaging data were acquired on a 3.0 T MRI scanner (Achieva TX, Philips Medical Systems) and a standard 32-channel radio-frequency head coil (Philips Medical Systems, Best, the Netherlands) at Hokkaido University. The IA patients in the second dataset were scanned twice, before and three months after initiation of therapy. T2*-weighted images were acquired with an echo-planar imaging (EPI) sequence for 6 minutes. The parameters used were as follows: repetition time (TR) 3,000 ms, echo time (TE) 30 ms, flip angle 80°, field of view 24 cm × 24 cm, matrix size 64 × 64, slice thickness 3.3 mm, interslice gap 3.3 mm, 48 axial slices, 120 volumes. During the scanning, the patients were instructed to rest calmly with their eyes open, not to sleep, and not to undergo any cognitive task. The structural T1 magnetization-prepared rapid gradient echo (MPRAGE) images of the head were also acquired with the following parameters: TR 7 ms, TE 3 ms, flip angle 8°, field of view 24 cm × 24 cm, matrix size 256 × 256, slice thickness 1.2 mm, interslice gap 1.2 mm, 170 sagittal slices.

### Data preprocessing

Preprocessing was performed using Statistical Parametric Mapping 12 software (SPM12; Wellcome Department of Cognitive Neurology, London, UK) and the CONN toolbox (www.nitrc.org/projects/conn) implemented in MATLAB (Mathworks, Natick, MA, USA). In the preprocessing pipeline, motion correction, realignment, slice-timing correction, outlier identification, coregistration to structural scan, segmentation, normalization to Montreal Neurological Institute (MNI) space, and spatial smoothing (8 mm Gaussian kernel) were performed. Structural scans were skull stripped, and segmented into grey matter, white matter and cerebrospinal fluid masks using the SPM 12 unified segmentation approach. For functional data, the four initial volumes were discarded for the magnetic field’s stabilization. Motion artifact was detected with the artifact detection toolbox (ART toolbox). Outliers’ images were added as nuisance regressors within the first-level general liner model (GLM). The noise was estimated and regressed out with CompCor, a component-based noise correction method. The white matter and cerebrospinal fluid masks were used as noise ROIs and regressed. A temporal band-pass filter of 0.008 to 0.09 Hz was applied to the time series for the removal of high-frequency activity associated with cardiac and respiratory activity. Residual BOLD time series were yielded to be extracted. A total of 132 atlas-based regions of interest (ROIs) from FSL Harvard-Oxford Atlas maximum likelihood cortical and subcortical atlas, and AAL atlas for cerebellum were used. For a detailed analysis of insular cortex (IC), we used probabilistic atlases of insular subregion, which included anterior IC consisting of anterior pole, anterior short gyrus, middle short gyrus and posterior short gyrus, and posterior IC consisting of anterior long gyrus and posterior long gyrus (Supplementary Table 3) [11].

### Dynamic functional connectivity analysis and clustering algorithm

Dynamic functional connectivity is a correlation coefficient of the windowed time series for characterizing temporal variability in functional connectivity. In the first level analysis, the sliding window approach was employed to estimate the windowed BOLD time series. The time course was segmented into 36-second windows, sliding the onset of each window by 18 seconds for a total of 18 windows. According to previous research, the selected duration of sliding windows would optimize the balance between capturing dynamic relationships and preserving reliable estimates of the correlated brain regional activity [18]. The fractions of last lengths of scans were round off. Fisher’s transformed correlation coefficient was computed in each sliding window between the windowed time course of the ROIs. In the second-level analysis, we initially evaluated recurrent connectivity patterns across IA patients in the dataset of first cohort (*n* = 33) as previously reported [10]. In brief, we first restructured each ROI-to-ROI matrix in one row (*Combination(132,2)* = 8646 dimensions) and window in the other dimension (18 rows). Second, we coupled all the matrices by the time dimension, producing a matrix of 8646 × 18 × 33 (number of patients) elements. Finally, the *k*-means++ cluster analysis algorithm was applied to the yielded matrix using cosine similarity metric. The clustering algorithm was applied 500 times to avoid local minimal value using a random initialization of centroid positions in each iteration. As a result, we acquired (i) *k* cluster centroids measured by cosine similarity (132 × 132 matrices), and (ii) a cluster label for the pattern corresponding to each matrix of the time window. Representative centroid matrices with functional connectivity values as recurrent patterns were calculated as averages of ROI-to-ROI matrices in each cluster pattern. For the second cohort’s dataset, we computed the cosine similarity between each functional connectivity matrix and the cluster centroids determined in the first dataset. Each matrix in the second dataset was then assigned the centroid label that minimized its distance with the centroids. Using the generated clustering, we calculated the occurrence probability of each cluster pattern for each individual. The top 10% ROI-to-ROI correlations within the absolute value of functional connectivity > 0.2 are rendered on the axial anatomical brain view, which were generated by BrainNet Viewer software [31].

### Statistical analysis

Analysis of covariance (ANCOVA) adjusting age and sex as confounds, or paired t-test was used to compare the unpaired or paired values of continuous variables, respectively. For multiple comparisons, we used Tukey-Kramer method to derive family-wise-error (FWE)-corrected p-value. Pearson product-moment correlation coefficient was used for a linear correlation. A receiver operating characteristic (ROC) analysis was performed to evaluate the accuracy for therapeutic responsiveness by functional connectivity value with the area under the curve (AUC).

To better understand the identified cluster patterns, differential functional connectivity in each cluster pattern was explored via the ROI-based inferences method. In brief, we defined a different set of connections for each row of the cluster-centroid matrix to classify all connections arising from the same ROI as a new set. We then performed a multivariate parametric GLM for all connections included in each of these new sets, generating an F-statistic for each ROI and a related uncorrected ROI-level p-value. Applying the Benjamini-Hochberg method, false discovery rate (FDR)-corrected ROI-level p-value was calculated as the expected proportion of false discoveries among all ROIs. In each statistically significant functional connectivity, we extracted the most differential functional connectivity with statistical significance in each cluster pattern by post-hoc Tukey-Kramer multiple comparison tests.

We used JMP Pro 14 (SAS Institute Inc., Cary, NC, USA) or MATLAB (Mathworks, Natick, MA, USA) for all analyses. The analysis results were considered to demonstrate statistical significance when the p-value including FDR- and FWE-corrected values was below 0.05. All statistical tests were two-sided.

## RESULTS

### Four distinct patterns of dynamic functional connectivity in patients with IA

We first partitioned the dynamic functional observations using the first dataset via *k*-means++ clustering, and the results enabled us to find distinct brain-wide coordination patterns (Figure 1A). The number of clusters *k* was determined through exploratory analyses varying *k* from 3 to 7 (Supplementary Figure 1A). Although a minimal intraclass correlation coefficient was obtained in the five cluster patterns (*k* = 5), the first pattern in cluster 5 was self-correlated with the other patterns from second to fifth (Supplementary Figure 1B). Even when the second to fifth pattern in cluster 5 were used, these modified four cluster patterns showed a lower intraclass correlation coefficient than the original four cluster patterns (Supplementary Figure 1C). Therefore, the number of clusters *k* = 4 (from the second to fifth pattern in the cluster *k* = 5) was adopted (Figure 1B). Among the cluster patterns, 5243 differentially correlated functional connections with statistical significance were detected (Figure 1C). We then applied the second dataset to the above separated clusters for validation.

**Figure 1.**
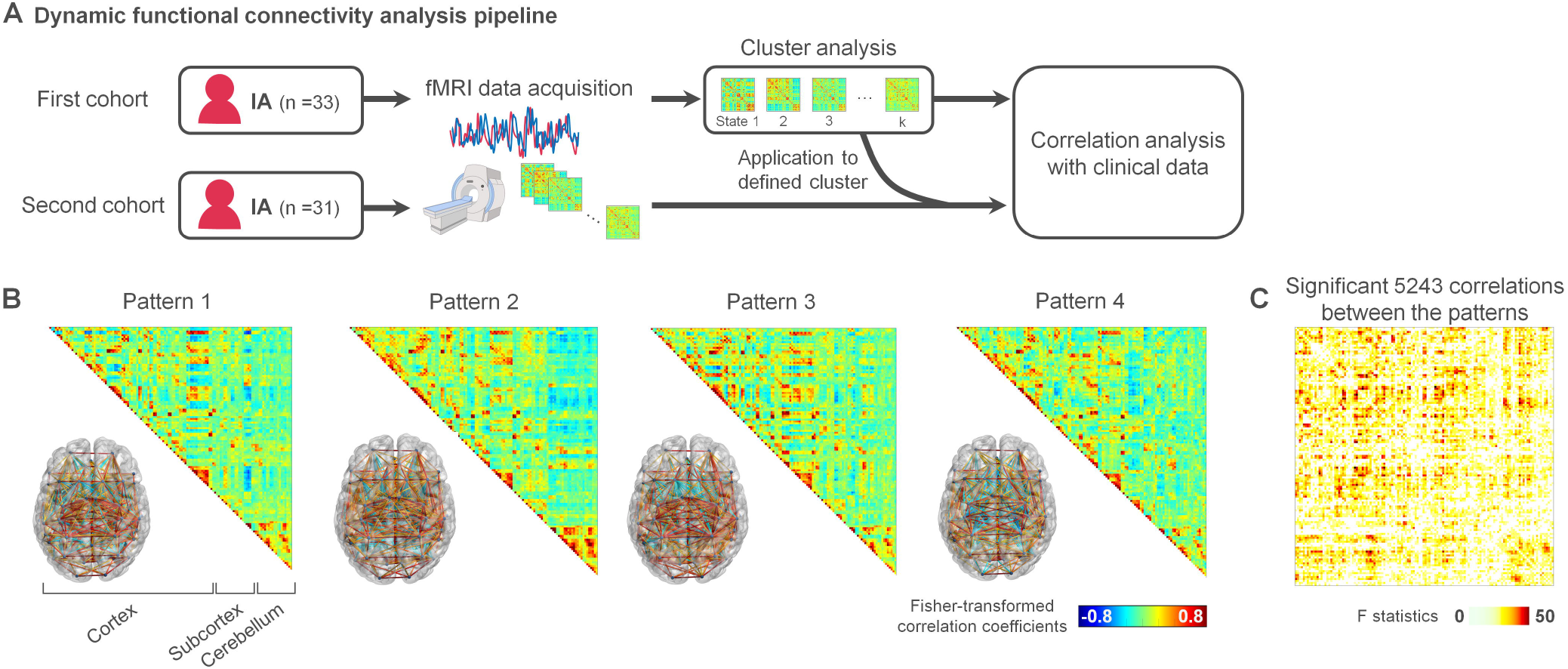
Distinct cluster patterns of dynamic functional connectivity in patients with inflammatory arthritis (IA). **(A)** Schema of dynamic functional connectivity analysis. (**B)** Four patterns by cluster analysis of dynamic functional connectivity matrices in the retrospective dataset with absolute connection value > 0.2 rendered on brain view. (**C**) Statistically significant functional connections colour-weighted by ANCOVA-F statics comparing the cluster patterns.

### Dynamic functional connectivity pattern in association with therapeutic response

The occurrence probability analysis of the connectivity patterns in the first dataset demonstrated that emerging pattern of cluster 4 was the least probable (Figure 2A), which was duplicated with the clusters of the second dataset (Figure 2B). In addition, in the first data set, the emergence rate of cluster 3 was higher in IA patients who responded favorably to biologics than those who did not (Figure 2C). In the second dataset, the occurrence of cluster 3 was more probable in the treatment-effective group than the treatment-ineffective group (Figure 2D), consistent with the results obtained from the first data set. The occurrence probability of cluster 3 was significantly associated with therapeutic responsiveness both in the first dataset with AUC 0.7462 (0.5771-0.9152) and in the prospective dataset with AUC 0.7083 (0.5224-0.8943) (Figure 2E, F). Furthermore, the occurrence probability of cluster 3 had a significant positive correlation with the therapeutic improvement rate of disease activity scoring by SDAI and ASDAS, and PGA (Figure 2G, H).

**Figure 2.**
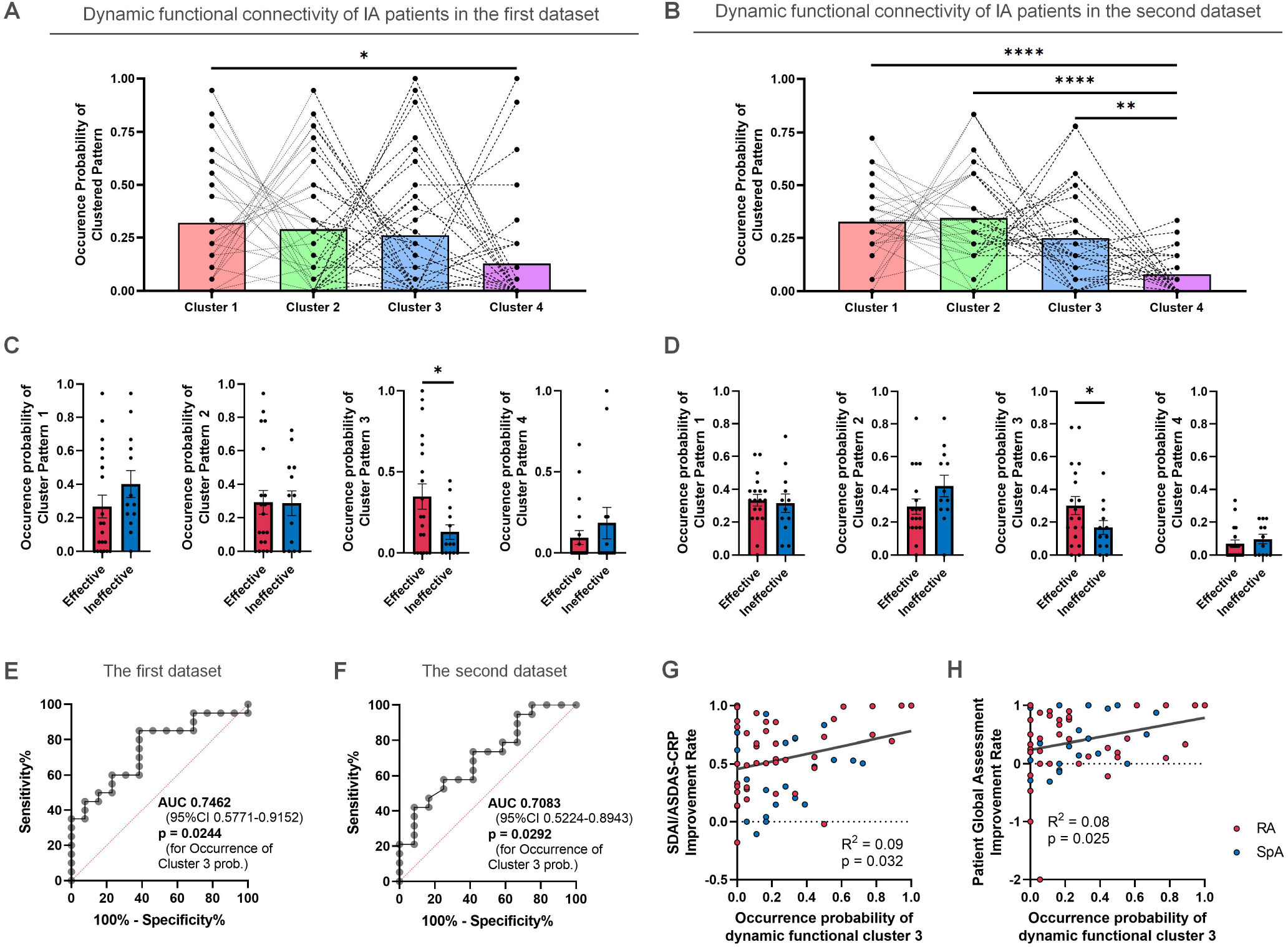
The occurrence probability of dynamic functional connectivity patterns related to therapeutic responsiveness in IA patients. (**A, B)** The likelihood of each coordination pattern occurrence associated with the patterns’ probability distribution in (**A**) the first dataset and (**B**) the second dataset. *P_FWE_ < 0.05, **P_FEW_ < 0.01, ****P_FWE_ < 0.0001, ANCOVA adjusting age and sex with post-hoc Tukey-Kramer test. (**C, D)** Comparison of occurrence probability of each pattern according to therapeutic effectiveness in (**C**) the first dataset and (**D**) the second dataset. *P < 0.05, ANCOVA adjusting age and sex. Data are mean ± s.e.m. (**E, F)** ROC analysis for therapeutic effectiveness using the occurrence probability of cluster pattern 3 in (**E**) the first and (**F**) second dataset. (**G, H)** Pearson’s correlation analysis between the occurrence probability of cluster pattern 3 and clinical parameters: **(G)** disease activity and **(H)** patients’ global assessment in the consolidated dataset.

### Dynamic corticocortical connectivity between left insular cortex and anterior cingulate cortex for therapeutic responsiveness

We examined what neurobiological features the cluster patterns represented, and detected that cluster 3 represents statistically higher functional correlations between corticocortical regions including subcortical areas, compared with the other cluster patterns (Figure 3A, B). On the other hand, the clusters 1, 2, and 4 represent the differential functional correlations in the frontoparietal cortex, cerebellum and subcortex itself, respectively (Figure 3C-E). Thus, the high probabilistic emergent pattern of corticocortical functional interaction was associated with therapeutic effectiveness in IA patients.

**Figure 3.**
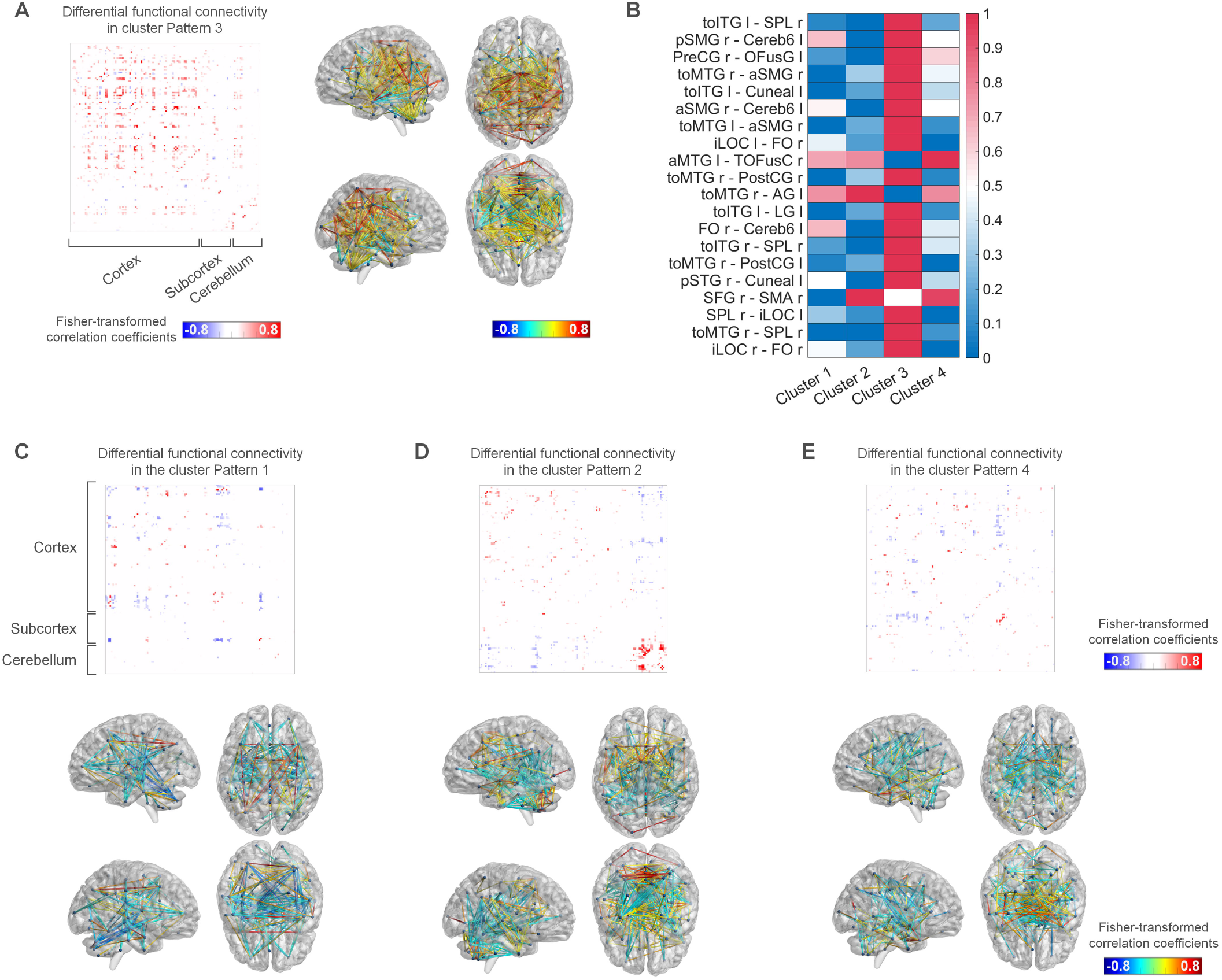
Therapeutic effectiveness in IA distinguished by dynamic functional corticocortical connectivity. (**A**) Statistically significant functional connectivity with high functional corticocortical correlations in cluster pattern 3 compared with other cluster patterns color-coded by F statics calculated from ANCOVA adjusting age and sex with Tukey-Kramer test. **(B)** Heatmap showing top 10 normalized values of functional connectivity between the regions of interest (ROIs) with statistical significance in cluster pattern 3. The abbreviations of ROIs are listed in Supplementary Table 5. **(C-E)** Statistically significant and differential functional connectivity with (**C**) frontoparietal cortex in cluster pattern 1, (**D**) cerebellum in the cluster pattern 2, and (**E**) subcortex itself in the cluster pattern 4 compared with the other cluster patterns color-coded by F statics calculated from ANCOVA adjusting age and sex with multiple comparisons using Tukey-Kramer test.

### Effect on the emergence probability of dynamic functional patterns by biologics

Finally, we analyzed the therapeutic effects on the emergence probability of dynamic functional connectivity clusters using the consolidated dataset (Supplementary Table 6). In the IA patients treated with tumor necrosis factor-α (TNF-α) inhibitors (*n* = 21), the occurrence probability of cluster pattern 3 was significantly higher in patients who responded favorably to biologic treatment than those who did not (Figure 4A). Also, the occurrence probability of cluster pattern 3 tended to be associated with therapeutic responsiveness in the IA patients treated with non-TNF-α inhibitors (*n* = 43) (Figure 4B). We analyzed whether therapeutic intervention itself affected the emergence probability of cluster 3 or not, using the fMRI data three months after biological therapy in the second dataset. It was decreased in the patients with therapeutic effectiveness while it was stable at low level in the patients with therapeutic ineffectiveness (Figure 4C).

**Figure 4.**
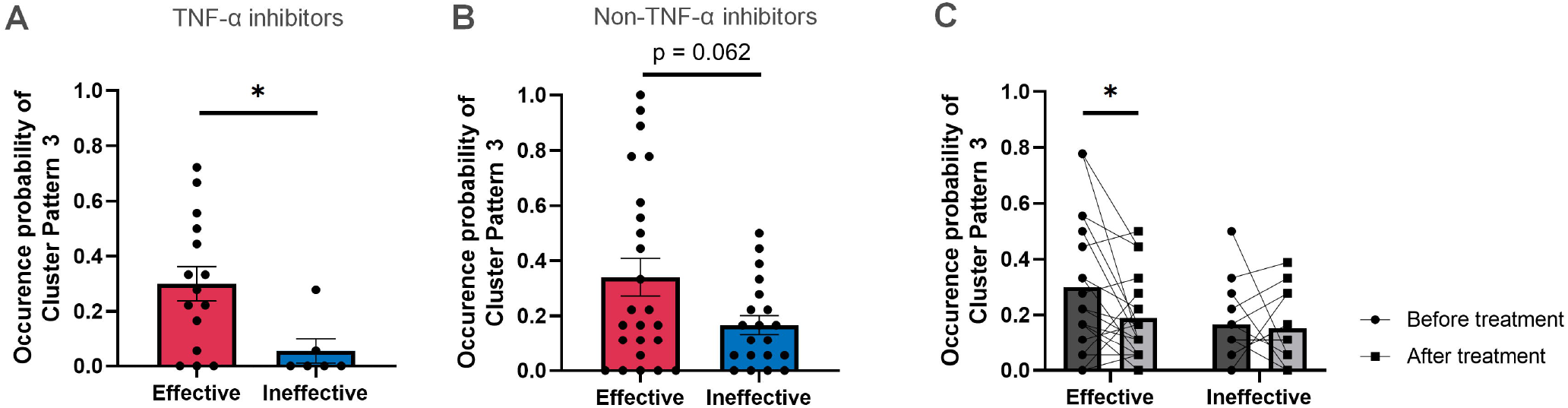
The occurrence probability of corticocortical connectivity patterns. (**A, B)** Occurrence probabilities of corticocortical cluster pattern 3 in IA patients treated with (**A**) TNF-α inhibitors (*n* = 21) or (**B**) non-TNF-α inhibitors (*n* = 43) in the consolidated dataset. *P < 0.05, ANCOVA adjusting age and sex. Data are mean ± s.e.m. (**C)** Occurrence probabilities of corticocortical cluster pattern 3 before and after initiation of biologics. fMRI data of the second dataset were used. *P < 0.05, paired t-test. Data are mean ± s.e.m.

## DISCUSSION

In this study, we aimed to study the brain’s dynamic organization, which determines patterns of functional connectivity associated with therapeutic responsiveness to biologics in IA patients. Results of our study identified that the dynamic patterns of long-standing functional correlations, especially high coherence of corticocortical connectivity, were associated with the improvement of disease activity and PGA by any biological treatments in IA patients. Furthermore, the occurrence probability of corticocortical dynamic pattern was reduced by biological treatments.

The dynamic states of whole-brain functional connectivity assessed by fMRI can demonstrate the flexibility of functional brain networks and capture the patterns of their transient alterations. The whole-brain patterns are widely associated with the physiopsychological status, indicating that distinct dynamic functional connectivity patterns would represent whole-brain functional configurations, which regulate the cognition, perception, and so on. Particularly, our study demonstrated that the occurrence probability of increased corticocortical functional correlations was related to treatment response in patients with IA. Cortical networks have been reported to be associated with pain processing via several mechanisms that represent and modulate pain. The cerebral cortex would regulate pain through interruption of noxious transmission from the spinal cord by activating the descending pain modulatory systems [23]. For instance, ACC, the insular cortex, the somatosensory cortices, the ventrolateral orbital cortex and the motor cortex have been proposed to be involved in the modulation of nociceptive processing. A previous report demonstrated decreased resting-state functional connectivity of the sensorimotor cortical network in patients with chronic pain [19]. Furthermore, occurrence probability of dynamic functional connectivity among brain cortical regions such as the prefrontal cortex, the posterior cingulate cortex and the angular gyrus, which construct default-mode network was positively associated with pain threshold [32]. In previous reports, biological treatments successfully reduced the activity of cortical regions including somatosensory cortex. Treatment with anti-TNF-α antibodies significantly decreased fMRI-assessed BOLD signals in the brain areas of RA patients within 24 hours [14]. Interestingly, responders to TNF-α antibodies showed significantly higher baseline brain activation in somatosensory cortex and association cortex than non-responders [25]. Similar response was also observed in Crohn’s disease [15]. As well as TNF-α, proinflammatory cytokines like IL-6 and IL-17 activate peripheral nociceptive afferents, sending inflammatory signals to the brain [28].

Based on these previous findings, a possible interpretation of our results is that the low coherence of resting-state corticocortical interactions would be associated with the dysfunction of modulating disability and pain, which may lead to less therapeutic efficacy such as insufficient improvement of disease activity regarding the subjective measure as PGA, although effective anti-rheumatic treatments have been shown to substantially reduce systemic inflammation levels. Proinflammatory cytokines would activate cortices to compensate the top-down controls. Results of the current study implied that anti-cytokine therapies could suppress corticocortical functional connectivity in the IA patients, which may partly account for the favorable responses observed. In other words, intrinsic structural and functional alterations of brain connectivity with the low coherence of resting-state corticocortical associations possibly lead to pain chronification and therapeutic unsatisfaction [22], as seen in the IA patients of our study, who did not satisfactorily respond to biological therapies. Not only systemic inflammation status but also brain functional status would be both important for the management of patients with IA. In IA patients with low cortical activation despite high disease activity, therapeutic approaches from the point of brain cortical function as well as anti-inflammatory treatments may be beneficial.

We acknowledge several limitations of this study. First, this study included a fairly small number of IA patients and consisted of a mixture of subjects with RA and SpA and treated with various biologics. Although the mixing analysis of diseases and therapeutic agents has a concern about the difference of underlying pathogenesis, it could simply demonstrate the functional alteration before and after anti-inflammatory therapy in patients with joint inflammation. Second, this study is of a retrospective nature. However, we were able to duplicate the findings from the first dataset with the second dataset, and we believe that the validated results strengthen the reliability of the findings obtained.

In conclusion, our study demonstrated that the dynamic pattern of functional connectivity was associated with therapeutic responsiveness to biologics in IA patients. Our data suggest that altered corticocortical interaction observed as low coherence of resting-state dynamic corticocortical functional connections would affect the clinical course of IA, and fMRI data of the brain as well as systemic inflammatory status may be of value to assess the therapeutic effect of biologics. Our study implies that brain function plays an important role in determining the course of IA and effectiveness of anti-rheumatic therapies.

## Data Availability

Data of fMRI are not publicly available and are stored locally, following national and Japanese laws on the protection of individuals with regard to the processing of personal data. Clinical data are available upon reasonable request. All data required to evaluate the conclusions are included in the article or uploaded as supplementary information.

## ACKNOWLEDGMENTS

We thank Naomi Tamura for the advice of statistical analysis, and Dr. Akito Tsutusmi for English editing.

## AUTHOR CONTRIBUTIONS

Conceptualization, Methodology, Software, Formal analysis and Writing – Original Draft: N.A. and K.S.; Conceptualization. Investigation, Writing – Original Draft and Visualization and Funding acquisition: Y.F.; Methodology, Software and Resources: K.K.T. and H.N.; Resources: K.K., M.Kono, and M.Kato; Software: M.Kanda; and Writing – Review & Editing, Supervision: T.A.;

## DECLARATION OF COMPTEING INTEREST

M.Kono has received research grants from Astellas, Chugai Pharmaceutical, GlaxoSmithKline, Hitachi, Kowa Company, Kyocera, Lotte, Mitsubishi Tanabe, Mochida, Nippon Shinyaku, Safofi, Taiju Life Social Welfare Foundation, Taisho Pharmaceutical, Takeda Pharmaceutical, Terumo Corporation and Yamazaki Baking, outside the submitted work; M.Kato has received research grants from AbbVie, Actelion, and GlaxoSmithKline, and speaking fees from Eli Lilly, outside the submitted work; T.A. has received research grants from Alexion, Astellas, AstraZeneca, Chugai Pharmaceutical, Daiichi Sankyo, Mitsubishi Tanabe, Otsuka Pharmaceutical, Pfizer and Takeda Pharmaceutical, and personal fees from AbbVie, Astellas, Boehringer Ingelheim, Bristol-Myers Squibb, Chugai Pharmaceutical, Daiichi Sankyo, Eisai, Eli Lilly, Medical & Biological Laboratories, Mitsubishi Tanabe, Novartis, Ono Pharmaceutical, Pfizer, Takeda Pharmaceutical, UCB Japan, outside the submitted work; and other authors declare no competing interests.

