## Supplementary for "Dynamics of corticocortical brain functional connectivity relevant to therapeutic response to biologics in inflammatory arthritis"

**This file includes:**

Supplementary Figures 1

Supplementary Tables 1 to 6

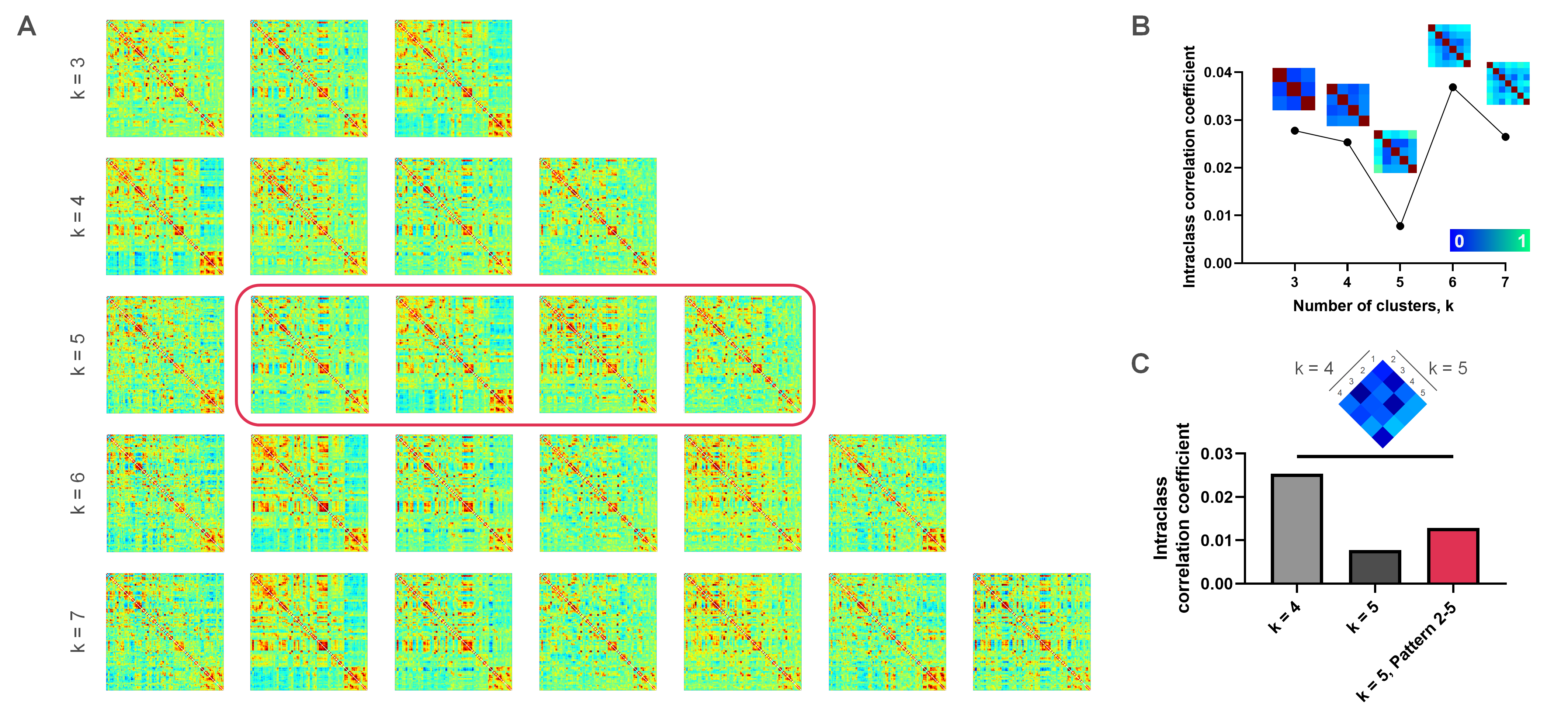
**Supplementary Figure 1. Exploration of dynamic coordination patterns.**

**(A)** Dynamic functional connectivity (FC) patterns obtained from the retrospective dataset using *k* = 3 to 7 number of clusters in the *k*-means++ clustering algorithm. (**B)** The minimal intraclass correlation coefficient of dynamic coordination pattern with *k* = 5 clusters. Matrices showed self-correlations of FC matrices in each cluster pattern. Strong self-correlation was found in the cluster number *k* = 5-7. (**C)** Even lower intraclass correlation coefficient of modified 4 cluster patterns using the second to fifth pattern in cluster number *k* = 5 than the patterns in cluster number *k* = 4. In correlation matrix, weak-correlation between four patterns in the cluster *k* = 4 and modified 2-5 patterns originally in the cluster *k* = 5.

| **Supplementary Table 1.** **Demographic and clinical characteristics in the first dataset.** | | | | | |
| --- | --- | --- | --- | --- | --- |
| **Characteristics** |  |  | IA (n = 33) | | |
|  |  |  | Before Treatment | | After Treatment |
| **Demographics** |  |  |  | |  |
| Age at fMRI, years |  |  | 62 [43-71.5] | |  |
| Sex, female |  |  | 25 (76%) | |  |
| PGA (0-10) |  |  | 7 [5-9] | | 3.2 [0.7-5.1] |
| Disease duration, years |  |  | 5 [0.5-15] | |  |
| **Disease classification** |  |  |  | |  |
| RA |  |  | 22 (67%) | |  |
| SpA |  |  | 11 (33%) | |  |
| **Laboratory findings** |  |  |  | |  |
| Rheumatoid factor |  |  | 20 (61%) | |  |
| ACPA |  |  | 19 (58%) | |  |
| C-reactive protein, mg/dL |  |  | 0.34 [0.05-2.02] | | 0.03 [0-0.135] |
| ESR, mm/h |  |  | 27 [16-56] | | 12 [8-26] |
| **Disease activity score** |  |  |  | |  |
| *for RA* |  |  | *(n = 22)* | | *(n = 22)* |
| TJC (0-28) |  |  | 6 [3-10] | | 1.5 [0-4] |
| SJC (0-28) |  |  | 4 [2-7] | | 0 [0-1] |
| EGA (0-10) |  |  | 4 [3-5] | | 1.5 [0.5-4] |
| SDAI |  |  | 21.8 [14.4-32.3] | | 7.1 [2.88-12.4] |
| *for SpA* |  |  | *(n = 11)* | | *(n = 11)* |
| ASDAS-CRP |  |  | 2.45 [2.15-3.40] | | 2.25 [1.69-2.64] |
| **Functional score** |  |  |  | |  |
| mHAQ score |  |  | 0.45 [0.175-1.5] | | 0.25 [0.05-1.1] |
| FIQR domain 1 |  |  | 7.3 [2.3-18.8] | | 5 [0.5-11.5] |
| FIQR domain 2 |  |  | 7 [2-13.5] | | 5 [2-8] |
| FIQR domain 3 |  |  | 19 [9.8-29.3] | | 14.5 [7-26] |
| FIQR total score |  |  | 33.8 [16.3-60.8] | | 25.3 [12.3-44.2] |
| **Therapeutic Regimen** |  |  |  | |  |
| Anti-TNF-α antibody |  |  | 12 (36%) | |  |
| Infliximab |  |  | 0 | |  |
| Adalimumab |  |  | 9 (27%) | |  |
| Etanercept |  |  | 0 | |  |
| Golimumab |  |  | 3 (9%) | |  |
| Certolizumab Pegol |  |  | 0 | |  |
| Anti-interleukin-6 antibody |  |  | 6 (18%) | |  |
| Abatacept |  |  | 5 (16%) | |  |
| Janus-kinase inhibitor |  |  | 6 (18%) | |  |
| Rituximab |  |  | 1 (6%) | |  |
| Anti-interleukin-17 antibody | |  | 2 (6%) | |  |
| Anti-interleukin-12/-23 antibody | |  | 0 | |  |
| **Concomitant Medication** | |  |  | |  |
| Glucocorticoid, mg/day | |  | 16 (48%), 5 [5-10] | |  |
| Methotrexate, mg/week | |  | 10 (42%), 9 [6-10.5] | |  |
| Salazosulfapyridine |  |  | 11 (46%) | |  |
| Tacrolimus |  |  | 5 (21%) | |  |
| Iguratimod |  |  | 4 (17%) | |  |
| Values are presented as n (%) or median (interquartile range). | | | |  | |
| Abbreviations: IA, inflammatory arthritis; fMRI, functional magnetic resonance imaging; PGA, patient global assessment; RA, rheumatoid arthritis; SpA, spondyloarthritis; ACPA, anti-citrullinated protein antibody; ESR, erythrocyte sedimentation rate; TJC, tenderness joint count; SJC, swollen joint count; EGA, evaluator global assessment; SDAI, simplified disease activity index; ASDAS, Ankylosing Spondylitis Disease Activity Score; mHAQ, modified Health Assessment Questionnaire; FIQR, Fibromyalgia Impact Questionnaire; TNF, tumor necrosis factor | | | | | |

| **Supplementary Table 2.** **Patients’ demographic and clinical characteristics in the second dataset.** | | | |
| --- | --- | --- | --- |
| **Characteristics** |  | IA (n = 31) | |
|  |  | Before Treatment | After Treatment |
| **Demographics** |  |  |  |
| Age at fMRI, years |  | 56 [45-68] | |
| Sex, female |  | 21 (68%) | |
| PGA (0-10) |  | 7 [5-9] | 4.5 [1.5-7] |
| Disease duration, years |  | 7 [1-12] | |
| **Disease classification** |  |  |  |
| RA |  | 19 (61%) | |
| SpA |  | 12 (39%) | |
| **Laboratory findings** |  |  |  |
| Rheumatoid factor |  | 15 (48%) | |
| ACPA |  | 15 (48%) | |
| C-reactive protein, mg/dL |  | 0.81 [0.04-3.1] | 0.03 [0-0.07] |
| ESR, mm/h |  | 25 [9-52] | 8 [5-14] |
| **Disease activity score** |  |  |  |
| *for RA* |  |  |  |
| TJC (0-28) |  | 4 [1-9] | 1 [0-2] |
| SJC (0-28) |  | 4 [0-6] | 0 [0-2] |
| EGA (0-10) |  | 6 [3-8] | 1.5 [0.5-4] |
| SDAI |  | 25.2 [11.2-33.3] | 7.1 [3-12] |
| *for SpA* |  |  |  |
| ASDAS-CRP |  | 3.11 [2.06-3.89] | 1.80 [1.06-2.99] |
| **Functional score** |  |  |  |
| mHAQ score |  | 0.55 [0.15-1.9] | 0.58 [0.01-0.89] |
| FIQR domain 1 |  | 14.7 [3.6-22] | 8.7 [2.1-17] |
| FIQR domain 2 |  | 10 [3.8-17] | 6 [2-11.8] |
| FIQR domain 3 |  | 27 [15.5-36.3] | 21.5 [9.9-32.4] |
| FIQR total score |  | 53.7 [26.3-74.3] | 33.5 [16.2-54.2] |
| **Therapeutic Regimen** |  |  |  |
| Anti-TNF-α antibody |  | 9 (29%) | |
| Infliximab |  | 1 (3%) | |
| Adalimumab |  | 3 (10%) | |
| Etanercept |  | 1 (3%) | |
| Golimumab |  | 0 | |
| Certolizumab Pegol |  | 3 (10%) | |
| Anti-interleukin-6 antibody |  | 11 (35%) | |
| Abatacept |  | 0 | |
| Janus-kinase inhibitor |  | 2 (6%) | |
| Rituximab |  | 2 (6%) | |
| Anti-interleukin-17 antibody |  | 5 (16%) | |
| Anti-interleukin-12/-23 antibody | | 1 (3%) | |
| **Concomitant Medication** |  |  |  |
| Glucocorticoid, mg/day |  | 9 (29%), 5 [4.5-6.5] | |
| Methotrexate, mg/week |  | 11 (33%), 10 [8-12] | |
| Salazosulfapyridine |  | 6 (19%) | |
| Tacrolimus |  | 1 (3%) | |
| Iguratimod |  | 0 | |
| Values are presented as n (%) or median (interquartile range). | | | |
| Abbreviations: IA, inflammatory arthritis; fMRI, functional magnetic resonance imaging; PGA, patient global assessment; RA, rheumatoid arthritis; SpA, spondyloarthritis; ACPA, anti-citrullinated protein antibody; ESR, erythrocyte sedimentation rate; TJC, tenderness joint count; SJC, swollen joint count; EGA, evaluator global assessment; SDAI, simplified disease activity index; ASDAS, Ankylosing Spondylitis Disease Activity Score; mHAQ, modified Health Assessment Questionnaire; FIQR, Fibromyalgia Impact Questionnaire; TNF, tumor necrosis factor | | | |

| **Supplementary Table 3.** **Baseline clinical parameters in IA patients of the first dataset.** | | | | |
| --- | --- | --- | --- | --- |
| **Characteristics** |  | IA (n = 33) | | |
|  |  | Treatment Effective (n = 20) | | Treatment Ineffective (n = 13) |
| **Demographics** |  |  | |  |
| Age at fMRI, years |  | 64 [55-72] | | 49 [36-69] |
| Sex, female |  | 15 (61%) | | 10 (39%) |
| PGA (0-10) |  | 5 [4-7.8] | | 7 [3.3-7] |
| Pain visual analogue scale (0-10) |  | 5 [4-7.8] | | 7 [3.3-7] |
| Disease duration, years |  | 5 [0-22] | | 4 [1.5-13] |
| **Disease classification** |  |  | |  |
| RA |  | 14 (70%) | | 8 (62%) |
| SpA |  | 6 (30%) | | 5 (38%) |
| **Laboratory findings** |  |  | |  |
| C-reactive protein, mg/dL |  | 0.53 [0.03-2.04] | | 0.28 [0.07-2.39] |
| ESR, mm/h |  | 27 [16-51] | | 30 [13-61] |
| **Disease activity score** |  |  | |  |
| *for RA* |  | *(n = 14)* | | *(n = 8)* |
| TJC (0-28) |  | 4 [3-6] | | 10 [6-13] |
| SJC (0-28) |  | 3 [1-5] | | 5 [2-8] |
| EGA (0-10) |  | 4 [3-5] | | 3.8 [2.3-6.5] |
| SDAI |  | 17.7 [13.4-26.4] | | 29.5 [14.8-34.8] |
| *for SpA* |  | *(n = 6)* | | *(n = 5)* |
| ASDAS-CRP |  | 2.43 [2.0-3.82] | | 2.64 [1.82-3.25] |
| **Functional score** |  |  | |  |
| mHAQ score |  | 0.38 [0.16-1.39] | | 0.75 [0.15-1.6] |
| FIQR domain 1 |  | 6.17 [1.83-18.1] | | 9.3 [5.67-19] |
| FIQR domain 2 |  | 2 [5-14] | | 10 [3-14] |
| FIQR domain 3 |  | 18 [9.1-32.9] | | 26.5 [14.8-29.3] |
| FIQR total score |  | 25.4 [13.1-59.0] | | 50.8 [19.8-60.8] |
| Values are presented as n (%) or median (interquartile range). | | |  | |
| Abbreviations: IA, inflammatory arthritis; fMRI, functional magnetic resonance imaging; PGA, patient global assessment; RA, rheumatoid arthritis; SpA, spondyloarthritis; ESR, erythrocyte sedimentation rate; TJC, tenderness joint count; SJC, swollen joint count; EGA, evaluator global assessment; SDAI, simplified disease activity index; ASDAS, Ankylosing Spondylitis Disease Activity Score; mHAQ, modified Health Assessment Questionnaire; FIQR, Fibromyalgia Impact Questionnaire | | | | |

| **Supplementary Table 4.** **Baseline clinical parameters in IA patients of the second dataset.** | | | | |
| --- | --- | --- | --- | --- |
| **Characteristics** |  | IA (n = 31) | | |
|  |  | Treatment Effective (n = 19) | | Treatment Ineffective (n = 12) |
| **Demographics** |  |  | |  |
| Age at fMRI, years |  | 54 [45-67] | | 47 [61-70] |
| Sex, female |  | 13 (68%) | | 8 (67%) |
| PGA (0-10) |  | 6 [5-8] | | 7.5 [5-9.8] |
| Pain visual analogue scale (0-10) |  | 7 [5-8] | | 7.5 [5-9.8] |
| Disease duration, years |  | 7 [1-12] | | 4 [1-17] |
| **Disease classification** |  |  | |  |
| RA |  | 12 (63%) | | 7 (58%) |
| SpA |  | 7 (37%) | | 5 (42%) |
| **Laboratory findings** |  |  | |  |
| C-reactive protein, mg/dL |  | 1.46 [0.04-4.52] | | 0.30 [0.04-2.04] |
| ESR, mm/h |  | 26 [8-64] | | 15 [10-49] |
| **Disease activity score** |  |  | |  |
| *for RA* |  | *(n = 12)* | | *(n = 7)* |
| TJC (0-28) |  | 6 [1-9] | | 2 [0-10] |
| SJC (0-28) |  | 4 [0-6] | | 2 [0-6] |
| EGA (0-10) |  | 5.5 [3-7] | | 6 [3-8] |
| SDAI |  | 25.5 [11.4-33.6] | | 18 [11.2-33.3] |
| *for SpA* |  | *(n = 7)* | | *(n = 5)* |
| ASDAS-CRP |  | 3.77 [2.11-3.96] | | 3.1 [1.94-3.46] |
| **Functional score** |  |  | |  |
| mHAQ score |  | 0.45 [0.04-1.9] | | 058 [0.05-1.7] |
| FIQR domain 1 |  | 13.7 [2.08-22.8] | | 15.3 [8.25-21.8] |
| FIQR domain 2 |  | 11 [2.8-17.3] | | 10 [5-14.8] |
| FIQR domain 3 |  | 27 [14.6-36.3] | | 28.5 [15.5-38.8] |
| FIQR total score |  | 55.1 [23.2-74.3] | | 52.8 [32.3-76.2] |
| Values are presented as n (%) or median (interquartile range). | | |  | |
| Abbreviations: IA, inflammatory arthritis; fMRI, functional magnetic resonance imaging; PGA, patient global assessment; RA, rheumatoid arthritis; SpA, spondyloarthritis; ESR, erythrocyte sedimentation rate; TJC, tenderness joint count; SJC, swollen joint count; EGA, evaluator global assessment; SDAI, simplified disease activity index; ASDAS, Ankylosing Spondylitis Disease Activity Score; mHAQ, modified Health Assessment Questionnaire; FIQR, Fibromyalgia Impact Questionnaire | | | | |

| **Supplementary Table 5.** **Centered MNI (Montreal Neurological Institute) coordinates of regions of interest (ROI).** | | | | |
| --- | --- | --- | --- | --- |
|  |  | MNI coordinates | | |
| Atlas | Brain Area | x | y | z |
| **FSL Harvard-Oxford Atlas maximum likelihood cortical atlas:** divided bilateral areas into left/right hemisphere (91 ROIs) | | | | |
|  | Frontal Pole, Right (FP r) | 26 | 52 | 8 |
|  | Frontal Pole, Left (FP l) | -25 | 53 | 8 |
|  | Insular Cortex, Right (IC r) | 37 | 3 | 0 |
|  | Insular Cortex, Left (IC l) | -36 | 1 | 0 |
|  | Superior Frontal Gyrus, Right (SFG r) | 15 | 18 | 57 |
|  | Superior Frontal Gyrus, Left (SFG l) | -14 | 19 | 56 |
|  | Middle Frontal Gyrus, Right (MidFG r) | 39 | 19 | 43 |
|  | Middle Frontal Gyrus, Left (MidFG l) | -38 | 18 | 42 |
|  | Inferior Frontal Gyrus, pars triangularis, Right (IFG tri r) | 52 | 28 | 8 |
|  | Inferior Frontal Gyrus, pars triangularis, Left (IFG tri l) | -50 | 28 | 9 |
|  | Inferior Frontal Gyrus, pars opercularis, Right (IFG oper r) | 52 | 15 | 16 |
|  | Inferior Frontal Gyrus, pars opercularis, Left (IFG oper l) | -51 | 15 | 15 |
|  | Precentral Gyrus, Right (PreCG r) | 35 | -11 | 50 |
|  | Precentral Gyrus, Left (PreCG l) | -34 | -12 | 49 |
|  | Temporal Pole, Right (TP r) | 41 | 13 | -30 |
|  | Temporal Pole, Left (TP l) | -40 | 11 | -30 |
|  | Superior Temporal Gyrus, anterior division, Right (aSTG r) | 58 | -1 | -10 |
|  | Superior Temporal Gyrus, anterior division, Left (aSTG l) | -56 | -4 | -8 |
|  | Superior Temporal Gyrus, posterior division, Right (pSTG r) | 61 | -24 | 2 |
|  | Superior Temporal Gyrus, posterior division, Left (pSTG l) | -62 | -29 | 4 |
|  | Middle Temporal Gyrus, anterior division, Right (aMTG r) | 58 | -2 | -25 |
|  | Middle Temporal Gyrus, anterior division, Left (aMTG l) | -57 | -4 | -22 |
|  | Middle Temporal Gyrus, posterior division, Right (pMTG r) | 61 | -23 | -12 |
|  | Middle Temporal Gyrus, posterior division, Left (pMTG l) | -61 | -27 | -11 |
|  | Middle Temporal Gyrus, temporooccipital part, Right (toMTG r) | 58 | -49 | 2 |
|  | Middle Temporal Gyrus, temporooccipital part, Left (toMTG l) | -58 | -53 | 1 |
|  | Inferior Temporal Gyrus, anterior division, Right (aITG r) | 46 | -2 | -41 |
|  | Inferior Temporal Gyrus, anterior division, Left (aITG l) | -48 | -5 | -39 |
|  | Inferior Temporal Gyrus, posterior division, Right (pITG r) | 53 | -23 | -28 |
|  | Inferior Temporal Gyrus, posterior division, Left (pITG l) | -53 | -28 | -26 |
|  | Inferior Temporal Gyrus, temporooccipital part, Right (toITG r) | 54 | -50 | -17 |
|  | Inferior Temporal Gyrus, temporooccipital part, Left (toITG l) | -52 | -53 | -17 |
|  | Postcentral Gyrus, Right (PostCG r) | 38 | -26 | 53 |
|  | Postcentral Gyrus, Left (PostCG l) | -38 | -28 | 52 |
|  | Superior Parietal Lobule, Right (SPL r) | 29 | -48 | 59 |
|  | Superior Parietal Lobule, Left (SPL l) | -29 | -49 | 57 |
|  | Supramarginal Gyrus, anterior division, Right (aSMG r) | 58 | -27 | 38 |
|  | Supramarginal Gyrus, anterior division, Left (aSMG l) | -57 | -33 | 37 |
|  | Supramarginal Gyrus, posterior division, Right (pSMG r) | 55 | -40 | 34 |
|  | Supramarginal Gyrus, posterior division, Left (pSMG l) | -55 | -46 | 33 |
|  | Angular Gyrus, Right (AG r) | 52 | -52 | 32 |
|  | Angular Gyrus, Left (AG l) | -50 | -56 | 30 |
|  | Lateral Occipital Cortex, superior division, Right (sLOC r) | 33 | -71 | 39 |
|  | Lateral Occipital Cortex, superior division, Left (sLOC l) | -32 | -73 | 38 |
|  | Lateral Occipital Cortex, inferior division, Right (iLOC r) | 46 | -74 | -2 |
|  | Lateral Occipital Cortex, inferior division, Left (iLOC l) | -45 | -76 | -2 |
|  | Intracalcarine Cortex, Right (ICC r) | 12 | -74 | 8 |
|  | Intracalcarine Cortex, Left (ICC l) | -10 | -75 | 8 |
|  | Frontal Medial Cortex (MedFC) | 0 | 43 | -19 |
|  | Juxtapositional Lobule Cortex, Right (SMA r) | 6 | -3 | 58 |
|  | Juxtapositional Lobule Cortex, Left (SMA l) | -5 | -3 | 56 |
|  | Subcallosal Cortex (SubCalC) | 0 | 21 | -15 |
|  | Paracingulate Gyrus, Right (PaCiG r) | 7 | 37 | 23 |
|  | Paracingulate Gyrus, Left (PaCiG l) | -6 | 37 | 21 |
|  | Cingulate Gyrus, anterior division (AC) | 1 | 18 | 24 |
|  | Cingulate Gyrus, posterior division (PC) | 1 | -37 | 30 |
|  | Precuneous Cortex (Precuneous) | 1 | -59 | 38 |
|  | Cuneal Cortex, Right (Cuneal r) | 9 | -79 | 28 |
|  | Cuneal Cortex, Left (Cuneal l) | -8 | -80 | 27 |
|  | Frontal Orbital Cortex, Right (FOrb r) | 29 | 23 | -16 |
|  | Frontal Orbital Cortex, Left (FOrb l) | -30 | 24 | -17 |
|  | Parahippocampal Gyrus, anterior division, Right (aPaHC r) | 22 | -8 | -30 |
|  | Parahippocampal Gyrus, anterior division, Left (aPaHC l) | -22 | -9 | -30 |
|  | Parahippocampal Gyrus, posterior division, Right (pPaHC r) | 23 | -31 | -17 |
|  | Parahippocampal Gyrus, posterior division, Left (pPaHC l) | -22 | -32 | -17 |
|  | Lingual Gyrus, Right (LG r) | 14 | -63 | -5 |
|  | Lingual Gyrus, Left (LG l) | -12 | -66 | -5 |
|  | Temporal Fusiform Cortex, anterior division, Right (aTFusC r) | 31 | -3 | -42 |
|  | Temporal Fusiform Cortex, anterior division, Left (aTFusC l) | -32 | -4 | -42 |
|  | Temporal Fusiform Cortex, posterior division, Right (pTFusC r) | 36 | -24 | -28 |
|  | Temporal Fusiform Cortex, posterior division, Left (pTFusC l) | -36 | -30 | -25 |
|  | Temporal Occipital Fusiform Cortex, Right (TOFusC r) | 35 | -50 | -17 |
|  | Temporal Occipital Fusiform Cortex, Left (TOFusC l) | -33 | -54 | -16 |
|  | Occipital Fusiform Gyrus, Right (OFusG r) | 27 | -75 | -12 |
|  | Occipital Fusiform Gyrus, Left (OFusG l) | -27 | -77 | -14 |
|  | Frontal Operculum Cortex, Right (FO r) | 41 | 19 | 5 |
|  | Frontal Operculum Cortex, Left (FO l) | -40 | 18 | 5 |
|  | Central Opercular Cortex, Right (CO r) | 49 | -6 | 11 |
|  | Central Opercular Cortex, Left (CO l) | -48 | -9 | 12 |
|  | Parietal Operculum Cortex, Right (PO r) | 49 | -28 | 22 |
|  | Parietal Operculum Cortex, Left (PO l) | -48 | -32 | 20 |
|  | Planum Polare, Right (PP r) | 48 | -4 | -7 |
|  | Planum Polare, Left (PP l) | -47 | -6 | -7 |
|  | Heschl''s Gyrus, Right (HG r) | 46 | -17 | 7 |
|  | Heschl''s Gyrus, Left (HG l) | -45 | -20 | 7 |
|  | Planum Temporale, Right (PT r) | 55 | -25 | 12 |
|  | Planum Temporale, Left (PT l) | -53 | -30 | 11 |
|  | Supracalcarine Cortex, Right (SCC r) | 8 | -74 | 14 |
|  | Supracalcarine Cortex, Left (SCC l) | -8 | -73 | 15 |
|  | Occipital Pole, Right (OP r) | 18 | -95 | 8 |
|  | Occipital Pole, Left (OP l) | -17 | -97 | 7 |
| **FSL Harvard-Oxford Atlas maximum likelihood subcortical atlas:** disregarded Cerebral White Matter, Cerebral Cortex, and Lateral Ventricular areas (15 ROIs) | | | | |
|  | Thalamus, Right (Thalamus r) | 11 | -18 | 7 |
|  | Thalamus, Left (Thalamus l) | -10 | -19 | 6 |
|  | Caudate, Right (Caudate r) | 13 | 10 | 10 |
|  | Caudate, Left (Caudate l) | -13 | 9 | 10 |
|  | Putamen, Right (Putamen r) | 25 | 2 | 0 |
|  | Putamen, Left (Putamen l) | -25 | 0 | 0 |
|  | Pallidum, Right (Pallidum r) | 20 | -4 | -1 |
|  | Pallidum, Left (Pallidum l) | -19 | -5 | -1 |
|  | Hippocampus, Right (Hippocampus r) | 26 | -21 | -14 |
|  | Hippocampus, Left (Hippocampus l) | -25 | -23 | -14 |
|  | Amygdala, Right (Amygdala r) | 23 | -4 | -18 |
|  | Amygdala, Left (Amygdala l) | -23 | -5 | -18 |
|  | Accumbens, Right (Accumbens r) | 9 | 12 | -7 |
|  | Accumbens, Left (Accumbens l) | -9 | 11 | -7 |
|  | Brain-Stem | 0 | -30 | -35 |
| **AAL Atlas:** Cerebellar parcellation (26 ROIs) | |  |  |  |
|  | Cerebellum Crus1, Left (Cereb1 l) | -36 | -66 | -30 |
|  | Cerebellum Crus1, Right (Cereb1 r) | 38 | -67 | -30 |
|  | Cerebellum Crus2, Left (Cereb2 l) | -29 | -73 | -38 |
|  | Cerebellum Crus2, Right (Cereb2 r) | 32 | -69 | -40 |
|  | Cerebellum 3, Left (Cereb3 l) | -9 | -37 | -19 |
|  | Cerebellum 3, Right (Cereb3 r) | 12 | -35 | -19 |
|  | Cerebellum 4 5, Left (Cereb45 l) | -14 | -44 | -17 |
|  | Cerebellum 4 5, Right (Cereb45 r) | 16 | -44 | -19 |
|  | Cerebellum 6, Left (Cereb6 l) | -23 | -58 | -24 |
|  | Cerebellum 6, Right (Cereb6 r) | 24 | -58 | -25 |
|  | Cerebellum 7b, Left (Cereb7 l) | -32 | -60 | -45 |
|  | Cerebellum 7b, Right (Cereb7 r) | 33 | -63 | -48 |
|  | Cerebellum 8, Left (Cereb8 l) | -26 | -55 | -48 |
|  | Cerebellum 8, Right (Cereb8 r) | 25 | -56 | -49 |
|  | Cerebellum 9, Left (Cereb9 l) | -11 | -49 | -46 |
|  | Cerebellum 9, Right (Cereb9 r) | 9 | -49 | -46 |
|  | Cerebellum 10, Left (Cereb10 l) | -23 | -34 | -42 |
|  | Cerebellum 10, Right (Cereb10 r) | 26 | -34 | -41 |
|  | Vermis 1 2 (Ver12) | 1 | -39 | -20 |
|  | Vermis 3 (Ver3) | 1 | -40 | -11 |
|  | Vermis 4 5 (Ver45) | 1 | -52 | -7 |
|  | Vermis 6 (Ver6) | 1 | -66 | -16 |
|  | Vermis 7 (Ver7) | 1 | -72 | -25 |
|  | Vermis 8 (Ver8) | 1 | -64 | -34 |
|  | Vermis 9 (Ver9) | 1 | -55 | -35 |
|  | Vermis 10 (Ver10) | 0 | -46 | -32 |
| **Probabilistic atlases of left insular subregions** | |  |  |  |
|  | Anterior Long Gyrus, Left | -37 | -12 | 6 |

| **Supplementary Table 6. Demographic and clinical characteristics in the consolidated dataset.** | | | | |
| --- | --- | --- | --- | --- |
| **Characteristics** |  |  | IA (n = 64) | |
|  |  |  | Treated by TNF-α inhibitors (n = 21) | Treated by non-TNF-α inhibitors (n = 43) |
| **Disease classification** |  |  |  |  |
| RA |  |  | 8 (38%) | 33 (77%) |
| SpA |  |  | 13 (62%) | 10 (23%) |
| **Therapeutic Regimen** |  |  |  |  |
| Anti-TNF-α antibody |  |  |  |  |
| Infliximab |  |  | 1 (5%) |  |
| Adalimumab |  |  | 12 (57%) |  |
| Etanercept |  |  | 1 (5%) |  |
| Golimumab |  |  | 4 (19%) |  |
| Certolizumab Pegol |  |  | 3 (14%) |  |
| Anti-interleukin-6 antibody |  |  |  | 18 (42%) |
| Abatacept |  |  |  | 6 (12%) |
| Janus-kinase inhibitor |  |  |  | 8 (19%) |
| Rituximab |  |  |  | 3 (6%) |
| Anti-interleukin-17 antibody | |  |  | 8 (19%) |
| Anti-interleukin-12/-23 antibody | |  |  | 1 (2%) |
| Values are presented as n (%) or median (interquartile range). | | | |  |
| Abbreviations: IA, inflammatory arthritis; RA, rheumatoid arthritis; SpA, spondyloarthritis; TNF, tumor necrosis factor | | | | |
